# Gene Discovery and Biological Insights into Anxiety Disorders from a Multi-Ancestry Genome-wide Association Study of >1.2 Million Participants

**DOI:** 10.1101/2024.02.14.24302836

**Authors:** Eleni Friligkou, Solveig Løkhammer, Brenda Cabrera-Mendoza, Jie Shen, Jun He, Giovanni Deiana, Mihaela Diana Zanoaga, Zeynep Asgel, Abigail Pilcher, Luciana Di Lascio, Ana Makharashvili, Dora Koller, Daniel S. Tylee, Gita A. Pathak, Renato Polimanti

**Author notes:** Correspondence: Renato Polimanti, PhD. Department of Psychiatry, Yale University School of Medicine. 60 Temple, Suite 7A, New Haven, CT 06511, USA.

## Abstract

We leveraged information from more than 1.2 million participants to investigate the genetics of anxiety disorders across five continental ancestral groups. Ancestry-specific and cross-ancestry genome-wide association studies identified 51 anxiety-associated loci, 39 of which are novel. Additionally, polygenic risk scores derived from individuals of European descent were associated with anxiety in African, Admixed-American, and East Asian groups. The heritability of anxiety was enriched for genes expressed in the limbic system, the cerebral cortex, the cerebellum, the metencephalon, the entorhinal cortex, and the brain stem. Transcriptome- and proteome-wide analyses highlighted 115 genes associated with anxiety through brain-specific and cross-tissue regulation. We also observed global and local genetic correlations with depression, schizophrenia, and bipolar disorder and putative causal relationships with several physical health conditions. Overall, this study expands the knowledge regarding the genetic risk and pathogenesis of anxiety disorders, highlighting the importance of investigating diverse populations and integrating multi-omics information.

## MAIN

Anxiety disorders affect many people worldwide, with a lifetime prevalence of approximately 34%, impacting overall health and all-cause mortality^1,2^. Although, according to the DSM-5^3^, this diagnostic category contains multiple anxiety disorders with distinct sets of diagnostic criteria, there is a significant phenotypic overlap among them; 48-68% of individuals with an anxiety diagnosis fulfill the criteria for at least a second one^4^. Likewise, twin and genome-wide association studies (GWAS) demonstrated a shared genetic basis for the whole spectrum of anxiety diagnoses^5^. Specifically, anxiety disorders have an estimated twin-based heritability of 20–60% across subtypes and are highly polygenic^6,7^. GWAS contributed to understanding the genetic architecture of anxiety disorders by identifying up to ten risk loci^8–10^. These were primarily conducted in individuals of European descent, with limited samples available from African American participants enrolled in the Million Veteran Program (MVP)^10^. While the lack of diversity is a known issue in human genetic research^11^, genetic research on anxiety appears to progress slower and be less diverse compared to current studies of other internalizing disorders, such as major depressive disorder (MDD) ^12^ and posttraumatic stress disorder (PTSD)^13^. To fill this gap, we conducted a multi-ancestry GWAS meta-analysis combining newly generated cohorts with previously reported data, reaching a total sample size of 1,266,780 participants (97,383 anxiety cases; Table 1). We included individuals of European (EUR, N=1,096,458), African (AFR, N=118,071), Admixed-American (AMR, N=36,634), South Asian (SAS, N=10,534), and East Asian (EAS, N=5,083) ancestries available from the All of Us Research Program (AoU)^14^, the Lundbeck Foundation Initiative for Integrative Psychiatric Research (iPSYCH)^15^, the FinnGen Project ^16^, MVP^10^, the Psychiatric Genomics Consortium (PGC)^17^, and the UK Biobank (UKB)^18^. Studying these cohorts, we identified multiple loci associated with anxiety and gained insights into the role of polygenic risk, pleiotropy, tissue-specific regulation, and transcriptomic and proteomic variations in the pathogenesis of anxiety disorders.

**Table 1.**
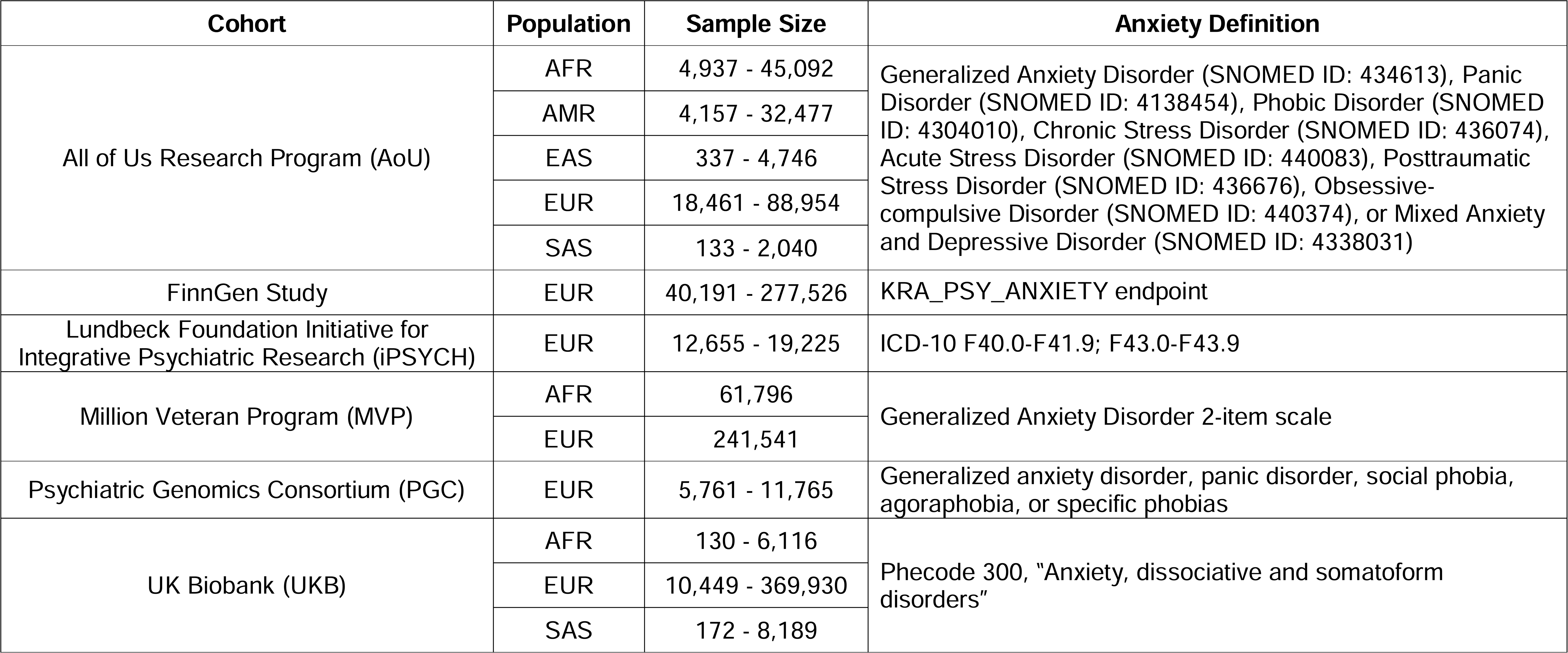
Cohorts and Anxiety Definitions investigated. Population groups included African (AFR), Admixed-American (AMR), East Asian (EAS), European (EUR), and South Asian (SAS) descent. Case and control sample sizes are reported for binary anxiety definitions, while the total sample size is reported for quantitative anxiety phenotypes. Other abbreviations include International Classification of Diseases (ICD) and Systemized Nomenclature of Medicine (SNOMED).

## RESULTS

### Genetic Discovery for Common Anxiety Factor, SNP-Heritability, and Genetic Correlation

We combined GWAS of anxiety from six cohorts consisting of five ancestries (Table 1). In the EUR cohort, statistically significant single nucleotide polymorphism (SNP)-based heritability was observed in all cohorts, with Z-scores ranging from 2.64 (PGC) to 15.13 (FinnGen), and values proportional to the effective sample size of each cohort. The genetic correlation (rg) ranged from 0.46 (p=2.1×10^-3^), between PGC and iPSYCH, to 1.02 (p=2.0×10^-5^), between PGC and AoU, with a median estimate of 0.72 (Figure 1; Supplemental Table 1). To quantify the genetically inferred differences across anxiety phenotypes across EUR datasets (Supplemental Figure 1), we applied PheMED (Phenotypic Measurement of Effective Dilution)^14^. FinnGen was considered the reference sample because it had the highest SNP-based heritability z score among the datasets investigated. MVP exhibited the largest phenotypic dilution (φ=2.92, p=5×10^-12^), followed by AoU (φ=2.35, p=6.0×10^-13^), PGC (φ=1.81, p=2.0-×10^-13^) and UKB (φ=1.73, p<1x10^-300^), while the iPSYCH anxiety phenotype was not significantly diluted when compared to FinnGen (φ=1.09, p=0.09). Leveraging data from cohorts assessed with different approaches, we observed that FinnGen and iPSYCH show more consistent genetic effects than the other samples investigated, with MVP showing the highest phenotypic dilution factor. We did not observe statistically significant SNP-based heritability among non-EUR groups due to their limited sample size (Supplemental Table 2). Therefore, we could not explore the genetic correlation and phenotypic dilution among anxiety datasets in other populations.

**Figure 1.**
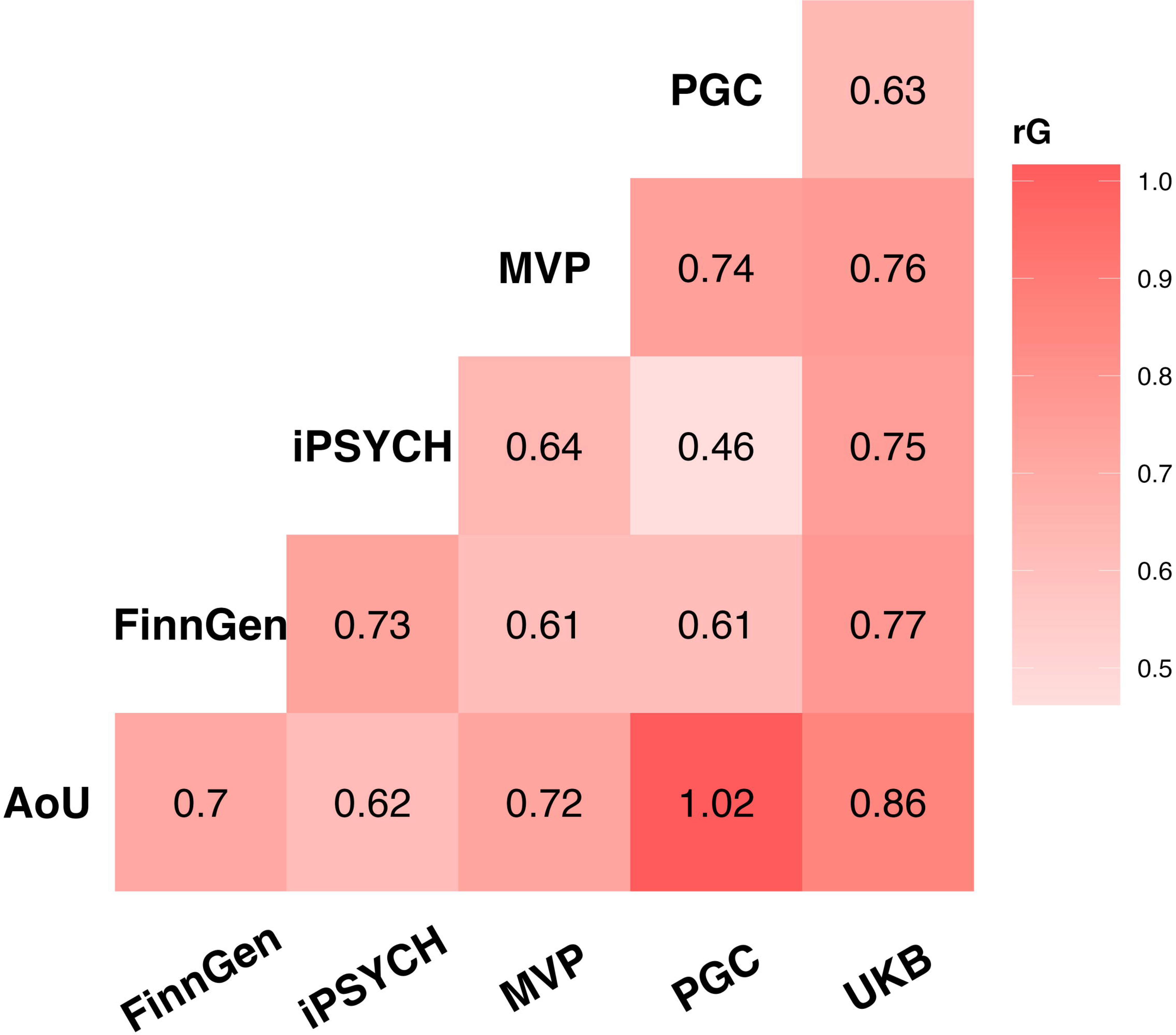
Genetic correlations among anxiety phenotypes assessed in participants of European descent (EUR). Full statistics are reported in Supplemental Table 1.

We combined the genome-wide information of the 1,096,458 EUR individuals with genomic structural equation modeling (gSEM)^15^ to account for the variability in the genetic correlation and the phenotypic heterogeneity among cohorts. The common anxiety factor (ANX; χ^2^(9)L=L13.4, p=0.15, comparative fit index=1, standardized root mean square residual=L0.08) loaded significantly on all indicators (standardized loadings on AoU=0.85±0.05, FinnGen=L0.82±0.03, MVP=0.80±0.04, iPSYCH=0.80±0.05, PGC=0.80±0.8, UKB=0.95±0.05; Figure 2). ANX showed a SNP-based heritability of 0.05±0.002 z-score=25.1) with no evidence of systematic bias due to population stratification or other confounders (linkage disequilibrium score regression^16^ LDSC intercept=1.01±0.01, ratio=0.01±0.02). We identified 35 linkage disequilibrium (LD)-independent (r^2^<0.1) variants with genome-wide significant association (p<5×10^-8^) with ANX (Figure 3; Supplemental Table 3). A conditional analysis^17^ yielded five additional genome-wide significant (GWS) variants, leading to a total of 40 genome-wide significant independent SNPs (Supplemental Table 3). Of these, 29 were LD-independent (LD r^2^<0.1) from variants reported by previous anxiety GWAS studies.^8–10^ Among the novel associations, the most significant one was rs6689226 (beta=0.01, p=4.8×10^-12^), located in an intergenic region downstream of *LINC01360* gene on chromosome 1.

**Figure 2.**
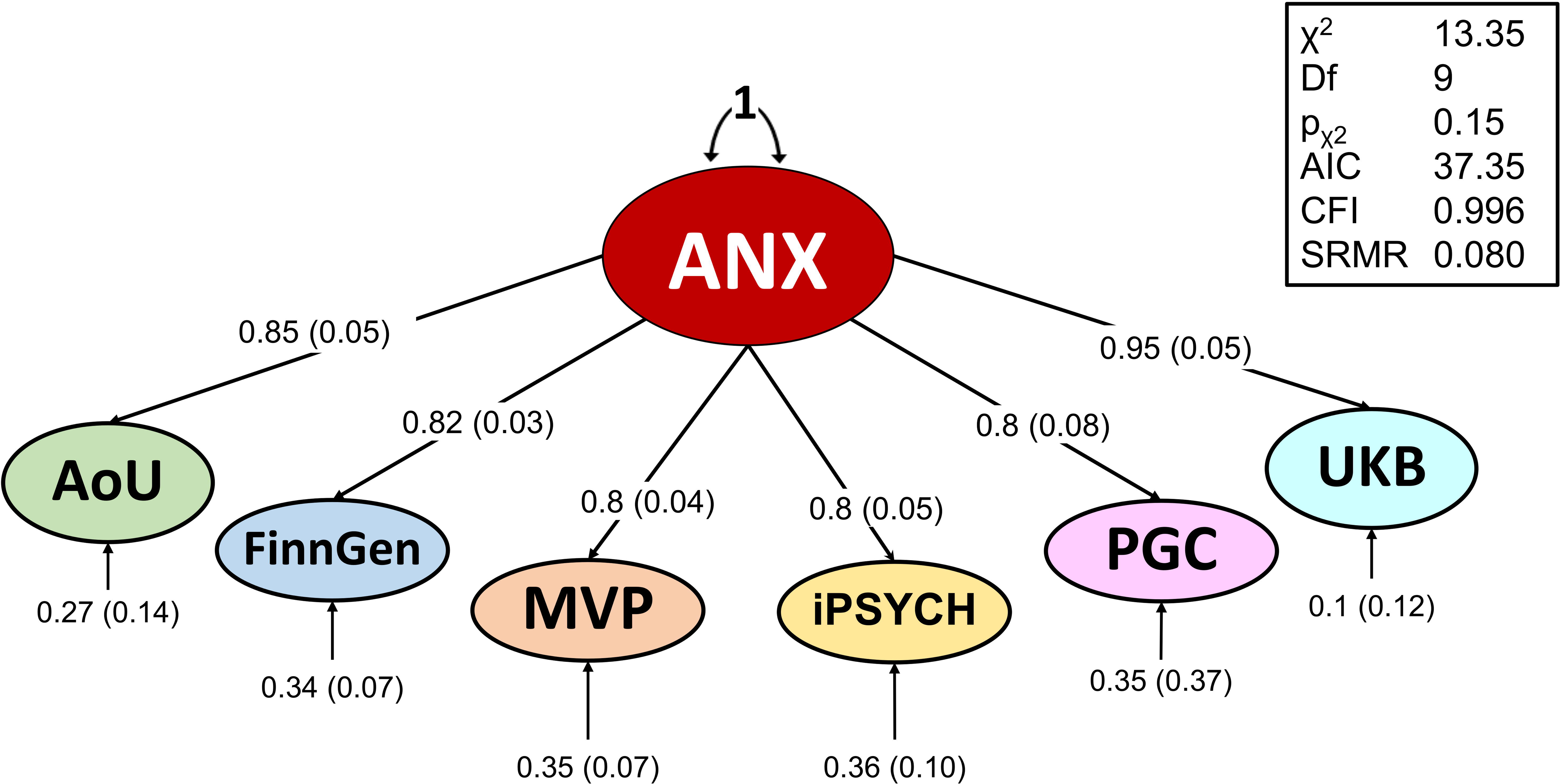
Factor structure of anxiety phenotypes assessed in individuals of European descent. Factor loadings and model fit for the confirmatory factor analysis model of the common anxiety factor (ANX) are reported.

**Figure 3.**
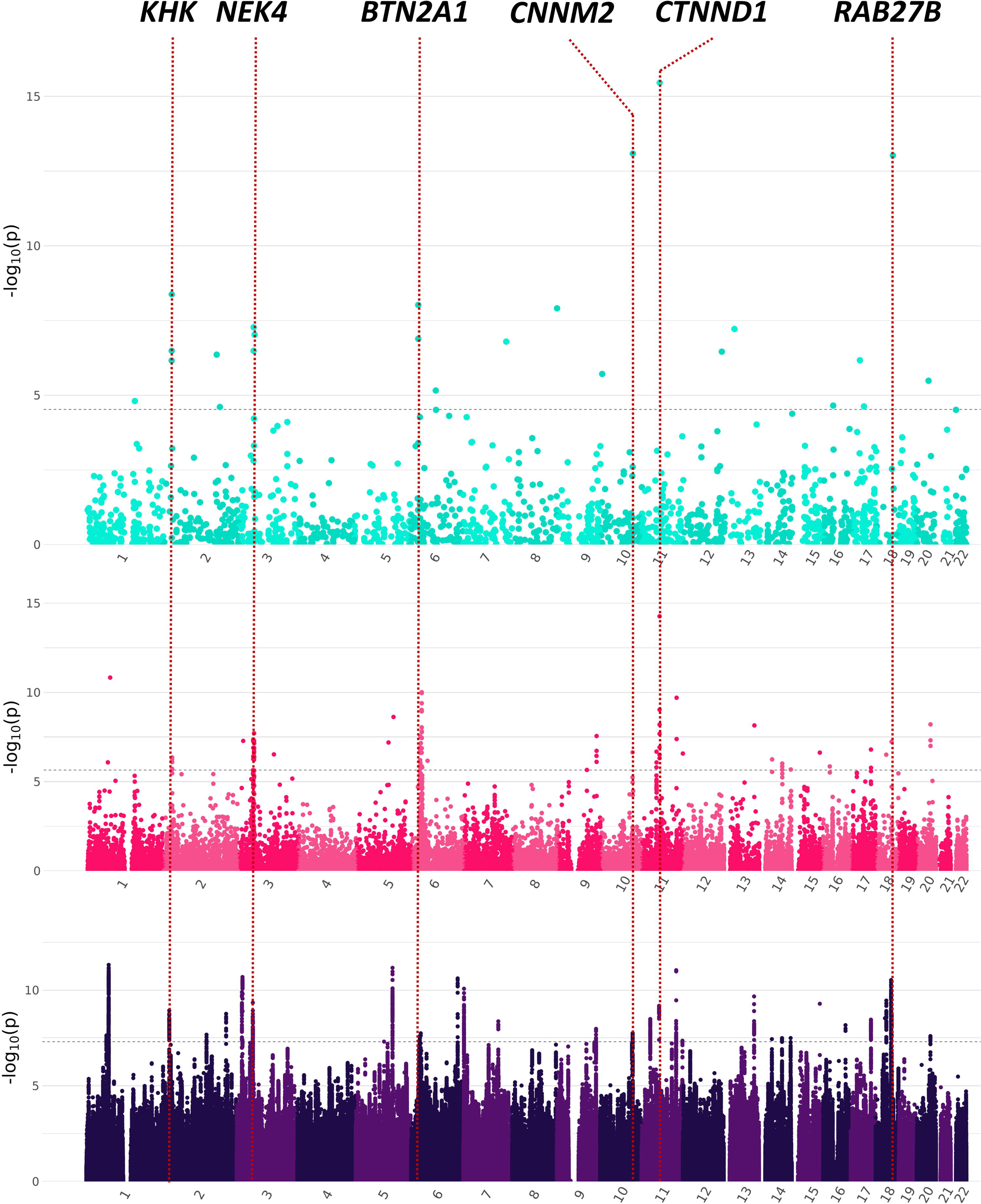
Manhattan plots of genome-wide, transcriptome-wide, and proteome-wide association statistics (bottom, center, and top, respectively) related to the common anxiety factor (ANX). Transcriptome-wide data are those obtained from the multiple-tissue analysis (Supplemental Table 8). Dashed lines represent Bonferroni multiple testing correction applied in each analysis. The labeled genes are those with convergent evidence across analyses.

Due to the limited sample size of other population groups, we could not apply the gSEM approach. These ancestry-specific meta-analyses were conducted using a sample-size weighted approach^18^ to combine genetic effects derived from different assessments (Table 1). In the AFR GWAS meta-analysis of UKB, MVP, and AoU, we observed a GWS association for rs575403075 (beta=-0.11, p=2.8×10^-8^), located within a candidate cis-regulatory element with enhancer-like signature (ENCODE accession number: EH38E2597848) on chromosome 7^19^. This AFR-specific locus (non-AFR minor allele frequency<0.01) was previously associated with anxiety in the AFR GWAS in MVP.^10^ No GWS associations were observed for the other populations. Because of the limited sample size of the other population groups, we observed cross-ancestry replication (p<0.01) only for two EUR GWS associations (Supplemental Table 3): rs12457101 (EUR beta=0.01, p=2.1×10^-9^; SAS beta=0.12, p=0.008) and rs10078721 (EUR beta=0.01, p=4.9×10^-8;^ AFR beta=0.02, p=0.009). EUR-derived polygenic risk scores (PRS) were significantly associated with anxiety phenotypes (fifth quintile vs. first quintile of PRS distribution) in AFR-AoU (OR=1.23, 95%CI=1.12-1.36), AMR-AoU (OR=1.55, 95%CI=1.39-1.74), EAS-AoU (OR=1.69, 95%CI=1.16-2.49), and EUR-AoU (OR=1.79, 95%CI=1.69-1.89) (Figure 4; Supplemental Table 4). The strength of the cross-ancestry PRS associations was proportional to the genetic distance between EUR and the other populations investigated^20^. Likely due to the limited sample size, null EUR-derived PRS association was observed with respect to SAS-AoU (Figure 4; Supplemental Table 4).

**Figure 4.**
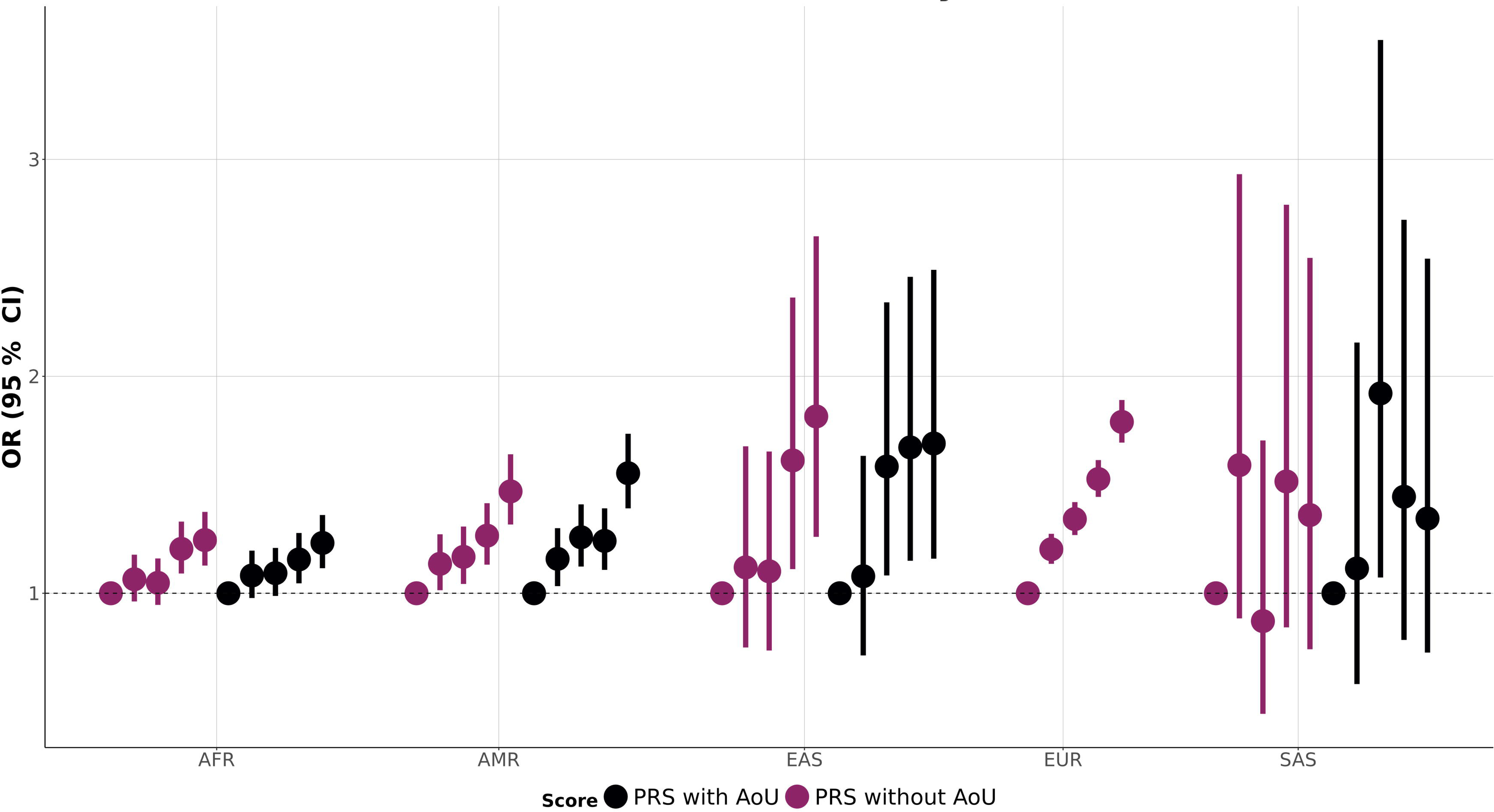
Within-ancestry and cross-ancestry polygenic risk score (PRS) associations of the common anxiety factor (ANX). Full statistics are available in Supplemental Table 4. In the within-ancestry, the EUR-ANX PRS (excluding EUR AoU from the training sample) is tested against the EUR AoU sample. In the cross-ancestry analysis, EUR-ANX PRSs (with and without EUR AoU in the training sample) are tested against AoU samples of African (AoU), Admixed-American (AMR), East Asian (EAS), and South Asian (SAS) descent.

A cross-ancestry meta-analysis revealed 41 GWS LD-independent loci, of which ten were novel (Supplemental Table 5), considering previously reported variants^8–10^ and index variants identified in the present ancestry-specific GWAS. The most significant among the novel cross-ancestry GWS associations was rs2510682 (cross-ancestry z=5.92, p=3.3×10^-9^; EUR beta=-0.12, p=6.2×10^-8^; AFR beta=-0.02, p=0.05; AMR beta=-0.04, p=0.18; EAS beta=-0.04, p=0.69; SAS beta=-0.04, p=0.44) located in an intronic region of *CNTN5* gene. For this locus and most of the other cross-ancestry associations, the signal was almost exclusively driven by the EUR sample. However, rs11681562 showed a cross-ancestry GWS association (z=-5.67, p=1.4×10^-8^) driven by both EUR and AFR samples (EUR beta=-0.01, p=4.3×10^-7^; AFR beta=-0.02, p=0.005; EAS beta=-0.05, p=0.64; SAS beta=-0.07, p=0.12).

### Transcriptomic and Proteomic Analyses

Partitioning the EUR-ANX heritability to 205 annotations based on tissue- and cell-specific transcriptomic profiles (Supplemental Table 6), we identified that ANX genetic liability was enriched for genes expressed in multiple brain regions, including the limbic system (p=2.3×10^-5^), the cerebral cortex (p=4.2×10^-5^), the cerebellum (p=7.4×10^-5^), the metencephalon (p=1.1×10^-4^), the entorhinal cortex (p=1.5×10^-4^), and the brain stem (p=2.0×10^-4^).

We performed a transcriptome-wide association study (TWAS) of the EUR-ANX GWAS using S-PrediXcan^21^. Initially, we conducted a tissue-specific TWAS considering 13 brain tissues available from GTEx v8^22^. We observed 152 transcriptome-wide significant associations related to 39 genes accounting for the number of genes and brain tissues tested (N=165,710, p<3.0×10^-7^) with a Bonferroni correction (Supplemental Table 7). The strongest association was related to the genetically regulated transcriptomic variation of the *DRD2* locus in the cerebellar hemisphere (z=-6.8, p=1.1×10^-11^). Interestingly, no associations survived Bonferroni correction for this gene among the other brain tissues tested. Conversely, four genes showed Bonferroni significant associations with 10 or more brain tissues: *CD40* (from hippocampus z=-6.0, p=1.5×10^-9^ to nucleus accumbens z=-5.1, p=2.7×10^-7^), *GNL3* (from spinal cord z=5.5, p=4.1×10^-8^ to cerebellum z=5.3, p=1.0×10^-7^), *NEK4* (from cerebellar hemisphere z=5.6, p=5.6×10^-8^ to spinal cord z=5.2, p=2.5×10^-7^), and *SMIM4* (from anterior cingulate cortex z=-5.9, p=3.4×10^-9^ to spinal cord z=-5.8, p=6.3×10^-9^). To investigate cross-tissue transcriptomic regulation further, we combined information from 49 tissues available from GTEx V8^22^ in a multi-tissue TWAS using S-MultiXcan^23^. After Bonferroni correction accounting for the number of genes tested (N=22,045; p<2.3×10^-6^), we identified 94 loci with genetically regulated transcriptomic associations with ANX (Figure 3, Supplemental Table 8). Fifty-nine of the 94 loci were not observed in the brain-specific TWAS, including the most significant transcriptome-wide association related to *MED19* (S-MultiXcan p=5.5×10^-15^). The second strongest multi-tissue association was *LINC01360* (S-MultiXcan p=1.5×10^-11^) near rs6689226, the most significant novel variant in the present EUR-ANX GWAS.

To understand the potential role of brain proteomic regulation in the pathogenesis of anxiety, we tested the EUR-ANX GWAS against protein quantitative trait loci (pQTL) in the dorsolateral prefrontal cortex (dlPFC) with FUSION^24^. We identified 24 loci with evidence of genetically regulated proteomic association with ANX after Bonferroni correction accounting for the number of genes tested (N=1,629; p<3.1×10^-5^) (Figure 3; Supplemental Table 9). We observed six genes identified by the proteome-wide association study (PWAS) to also be Bonferroni significant in the multi-tissue TWAS: *CTNND1* (PWAS p=3.5×10^-16^; multi-tissue TWAS p=2.1×10^-8^), *CNNM2* (PWAS p=8.3×10^-14^; multi-tissue TWAS p=2.3×10^-7^), *RAB27B* (PWAS p=9.4×10^-14^; multi-tissue TWAS p=5.9×10^-8^), *KHK* (PWAS p=4.2×10^-9^; multi-tissue TWAS p=8.1×10^-7^), NEK4 (PWAS p=9.4×10^-8^; multi-tissue TWAS p=7.2×10^-8^), and *BTN2A1* (PWAS p=9.6×10^-9^; multi-tissue TWAS p=1.2×10^-7^). Of those, *CTNND1* and *NEK4* also demonstrated Bonferroni-significant expression in areas of the cortex, the basal ganglia, and the cerebellum in the brain TWAS. *CGREF1* was a PWAS significant hit (PWAS p=3.3×10^-7^), with a significant brain TWAS expression in the frontal cortex (TWAS p=1.13×10^-7^) and the putamen (TWAS p=2.73×10^-7^). The Bonferroni-significant PWAS loci were enriched for multiple synaptic locations and biological processes, including synapse (gene ontology, GO:0045202, p=4.6×10^-4^), synaptic vesicle membrane (GO:0030672, p=5.9×10^-4^), presynapse, synaptic vesicle (GO:0008021, p=0.001), process in the synapse (SYNGO:synprocess, p=0.003), and process in the presynapse (SYNGO:presynprocess, p=0.008).

### Pleiotropy with Human Traits and Diseases

We used MixeR^25^ to investigate the polygenic architecture of anxiety in the context of other psychiatric disorders. A total of 12,622±834 influential variants were estimated for EUR-ANX. A similar estimate was observed for MDD (N=11,428±453, p_ANX-difference_=0.21). Conversely, for anorexia nervosa (AN, N=7,869±387, p_ANX-difference_=1.9×10^-6^), attention-deficit hyperactivity disorder (ADHD, N=7,978±387, p_ANX-difference_=4.4×10^-7^), bipolar disorder (BIP, N=8,772±387, p_ANX-difference_=1.4×10^-5^), PTSD (N=7,585±533, p_ANX-difference_=3.6×10^-7^), and schizophrenia (SCZ, N=9,636±262, p_ANX-difference_=6.4×10^-4^), the number of influential variants was significantly lower. No reliable estimate was obtained for autism spectrum disorder (ASD), obsessive-compulsive disorder (OCD), and Tourette syndrome (TS, Supplemental Table 10). Considering statistically meaningful bivariate MixeR models (Akaike information criterion for the best model versus model with minimal possible polygenic overlap>0; Supplemental Table 11), ANX shared 91±4%, 75±9%, and 68±1% of its influential variants with MDD, BIP, and SCZ, respectively (Figure 5A; Supplemental Figure 2).

**Figure 5.**
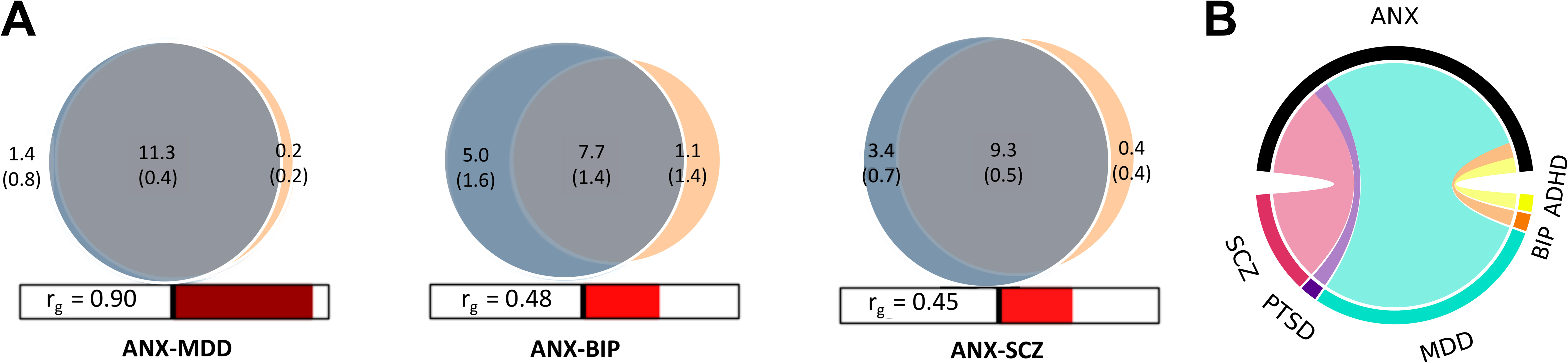
**A.** Venn diagrams of unique and shared causal variants showing polygenic overlap (gray) of the common anxiety factor (ANX) with major depressive disorder (MDD), bipolar disorder (BIP), and schizophrenia (SCZ). The numbers indicate the estimates of causal variants and their standard errors (in thousands), explaining 90% of the SNP heritability in each phenotype. The size of the circles reflects the degree of polygenicity. **B.** Chord diagram representing ANX local genetic correlations with respect to attention deficit and hyperactivity disorder (ADHD), BIP, MDD, post-traumatic stress disorder (PTSD), and SCZ.

To estimate the local genetic correlation of ANX with other psychiatric traits, we used Local Analysis of (co-)Variant Association (LAVA)^26^. After Bonferroni correction for the number of tests performed (N=12,368, p<4.1×10^-6^), we observed 47 local genetic correlations of ANX with five psychiatric disorders (i.e., ADHD, BIP, MDD, PTSD, and SCZ) across 35 genomic regions (Figure 5B; Supplemental Table 12). Consistent with the extensive genetic overlap observed in MixeR, we observed local genetic correlations between ANX and MDD in 39 genomic regions. Twelve regions showed evidence of local genetic correlation between ANX and SCZ. Only two regions were genetically correlated between ANX and ADHD, BIP, and PTSD, respectively. ANX demonstrated local genetic correlations with two or more psychiatric disorders in 10 regions. Among them, ANX showed local genetic correlation with MDD (rho=0.97, p=2.3×10^-6^), SCZ (rho=0.84, p=7.2×10^-12^), and BIP (rho=0.73, p=8.3×10^-8^) within locus 1719 (chr11:112,755,447-113,889,019; Supplemental Figure 3. Within locus 2281 (chr18:52,512,524-53,762,996), ANX local genetic correlation was found with MDD (rho=0.88, p=1.7×10^-12^) and SCZ (rho=0.67, p=1.9×10^-7^). Among the most significant genomic regions, we also observed locus 852 (chr5:91,956,906-93,814,604), where ANX was genetically correlated with ADHD (rho=0.97, p=4.7×10^-7^) and MDD (rho=1, p=3.4×10^-8^).

To decompose the pleiotropic mechanisms linking ANX to multiple psychiatric disorders, we applied LAVA multiple regression models^26^ to the five genomic regions with the strongest evidence of local genetic correlation with ANX (Supplemental Table 13). ANX was entered in the regression models as an outcome, and the other genetically correlated traits as predictors. We observed that the local genetic correlations of ANX with MDD, SCZ, and BIP in locus 1719 were not independent of each other. Shared pleiotropic mechanisms involving ANX were also observed in locus 62 (chr1:72,513,120-73,992,170) for MDD and SCZ and locus 852 (chr5:91,956,906-93,814,604) for ADHD and MDD. Conversely, for locus 1582 (chr10:106,142,284-107,877,787), we identified independent pleiotropic mechanisms of ANX with MDD (rho=0.91, p=5.2×10^-9^; gamma=0.63, p=0.04) and SCZ (rho=1, p=2.8×10^-8^; gamma=0.78, p=0.004). For locus 2281 (chr18:52,512,524-53,762,996), the ANX-MDD local genetic correlation (rho=0.88, p=1.7×10^-12^; gamma=0.78, p=0.003) was the primary pleiotropic driver that also accounted for ANX-SCZ relationship (rho=0.67, p=1.9×10^-7^; gamma=0.16, p=0.35).

To investigate the shared genetic mechanisms between ANX and human traits and diseases, we performed a phenome-wide genetic correlation analysis leveraging 11,175 phenotypes available from UKB, MVP, and FinnGen^27–29^. After Bonferroni correction (p<4.47×10^-6^), 1,929 showed a statistically significant genetic correlation with ANX (Figure 6, Supplemental Table 14). Among these, the median |rg| was 0.42 (25^th^ percentile=0.29; 75^th^ percentile=0.55), with 77 of the top 100 results (all |rg|>0.72) being mental health outcomes (e.g., FinnGen anxiety disorders rg=0.91, p<1x10^-300^; UKB self-reported depression rg=0.83, p=1.0×10^-53^) and related drug prescriptions (e.g., UKB citalopram medication rg=0.93, p=7.3×10^-8^; FinnGen depression medications rg=0.82, p<1x10^-300^). Among non-psychiatric outcomes, the top results included myalgia (FinnGen rg=0.85, p=4.0×10^-12^), nausea/vomiting (UKB rg=0.84, p=1.5×10^-11^), diseases of pulp and periapical tissues (FinnGen rg=0.84, p=4.6×10^-7^), dysuria (MVP rg=0.79, p=1.7×10^-6^), cystitis (FinnGen rg=0.77, p=8.1×10^-9^), constipation (UKB rg=0.75, p=1.2×10^-20^), pain related to temporomandibular disorders (FinnGen rg=0.75, p=1.4×10^-16^), unspecified dorsalgia (FinnGen rg=0.73, p=3.1×10^-45^), and pain management (UKB rg=0.73, p=6.1×10^-30^).

**Figure 6.**
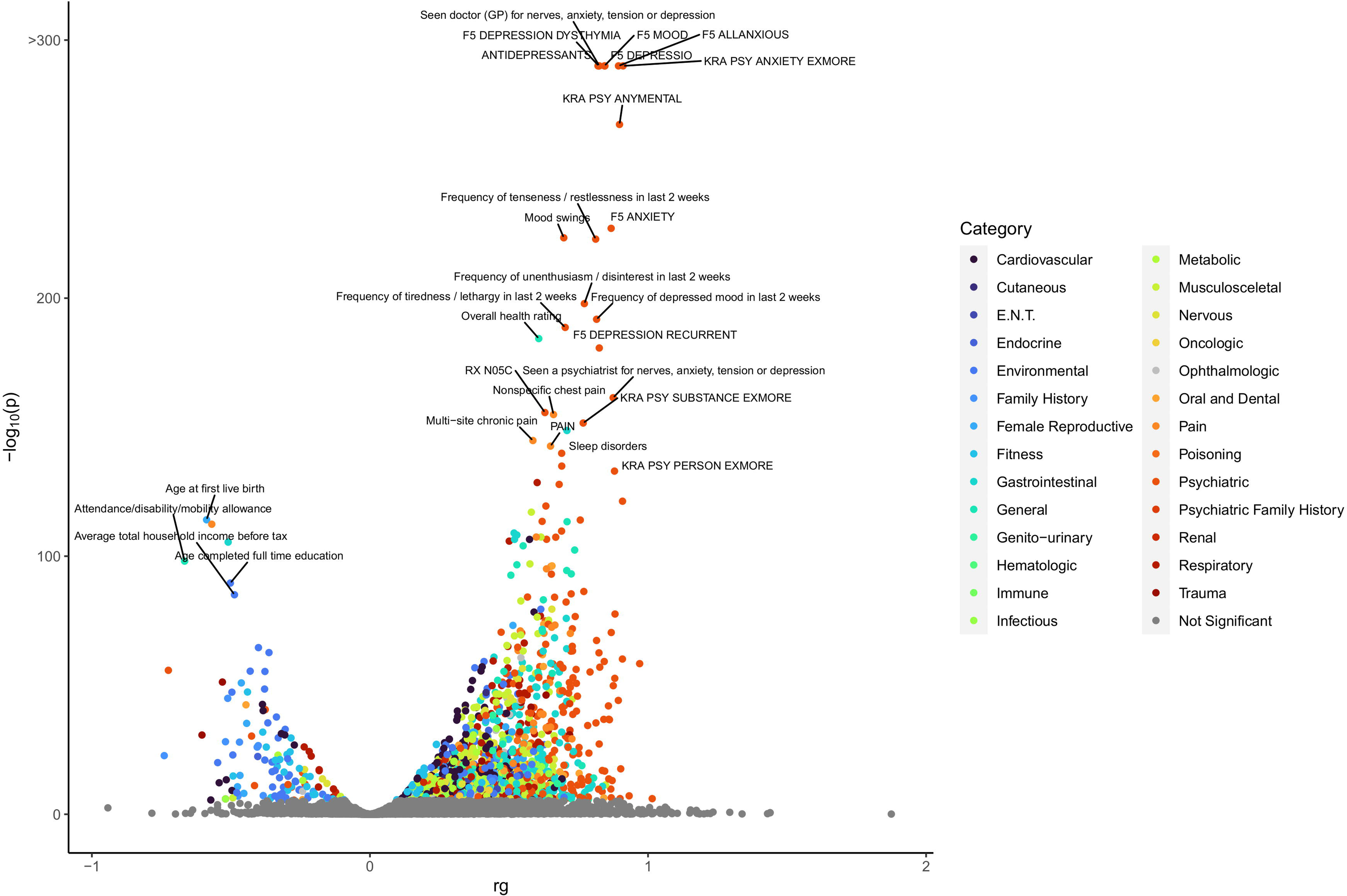
Phenome-wide genetic correlation of the common anxiety factor (ANX). The x-axis reports the genetic correlation of ANX with traits available from UK Biobank, FinnGen, and Million Veteran Program. The y-axis corresponds to two-tailed −log_10_(p-value). Bonferroni-significant results are color-coded based on the corresponding categories. Full statistics of the Bonferroni-significant results are available in Supplemental Table 14. Non-significant results are reported in grey.

To investigate potential causal relationships underlying those genetic correlations, we employed the latent causal variable approach (LCV)^30^. We identified 24 traits with statistically significant genetic causal proportion (gcp) after a false discovery rate (FDR) multiple testing correction (FDR q<0.05, p<4.1×10^-6^) (Supplemental Table 15). Positive causal effects were observed from other phenotypes to ANX in all cases (LCV rho>0). Only three of these phenotypes were related to mental health: “Number of things worried about during worst period of anxiety” (UKB gcp=0.89, p=4.1×10^-63^; rho=0.67±0.10), “Ever had period of mania/excitability” (UKB gcp=0.46, p=1.4×10^-28^; rho=0.51±0.08), and “Number of cigarettes currently smoked daily” (UKB gcp=0.54, p=2.2×10^-7^; rho=0.23±0.07). Seven of the phenotypes identified were related to physical health concerns, with the strongest relationship implicating Dupuytren’s disease (MVP, gcp=0.71, p=7.1×10^-93^; rho=0.13±0.05). Five were related to the cardiovascular system: “Atherosclerotic cardiovascular disease” (MVP, gcp=0.60, p=8.9×10^-29^; rho=0.40±0.09), “Other disorders of circulatory system” (UKB, gcp=0.43, p=5.2×10^-26^; rho=0.46±0.07), “Medication for transient ischemic attack” (MVP, gcp=0.63, p=6.4×10^-17^; rho=0.24±0.0.8), “Other specified cardiac dysrhythmias” (UKB, gcp=0.93, p=7.0×10^-15^; rho=0.40±0.08), and “Heart Surgery” (UKB, gcp=0.96, p=7.0×10^-15^; rho=0.40±0.0.08). We also observed two environmental variables with FDR-significant effects: “Workplace had a lot of diesel exhaust” (UKB, gcp=0.95, p=2.9×10^-24^; rho=0.45±0.10) and “Number of gap periods in employment” (UKB, gcp=0.60, p=5.4×10^-7^; rho=0.26±0.08).

## DISCUSSION

The present study analyzed genome-wide information of more than 1.2 million participants, giving valuable insights into the genetic liability of anxiety disorders. The ANX GWAS in EUR identified 40 GWS associations, 29 of which were novel. Unfortunately, the limited sample size of other ancestry groups permitted us to only confirm a previously reported locus in AFR^10^. However, the cross-ancestry GWAS added ten novel anxiety risk loci to our gene discovery. The novel findings of the EUR-specific and cross-ancestry analyses quadrupled the gene discoveries reported by previous studies^8–10^. Among the novel loci identified, the strongest ANX association was with rs6689226, located near *LINC01360*. While the regulatory mechanisms related to this long non-coding RNA (lncRNA) are unclear, this variant was previously associated with several mental health outcomes such as smoking initiation^31^, externalizing behaviors^32^, and cross-disorder psychopathology^33^. Our multi-tissue transcriptomic analysis identified genetically regulated expression of *LINC01360* in the testis. LncRNAs are more expressed in the testis than other tissues, and some of them play a role in androgen production^34^, which may be of pathophysiological relevance in certain animal models of anxiety^35^. In the cross-ancestry GWAS, the strongest ANX association was rs2510682, located in an intron region of the *CNTN5* gene, which encodes a protein mediating cell surface interactions during the development of the nervous system^36^. While it has not been associated with anxiety by previous GWAS^8–10^, *CNTN5* variants were linked to neuroticism^37^, suicidal behaviors^38^, and ASD^39^. The second most significant cross-ancestry association, rs11681562, was also a novel ANX locus. Variants in high LD with rs11681562 (r^2^>0.8) have been previously associated with multiple phenotypes related to human cortical folding (i.e., vertex-wise sulcal depth, vertex-wise cortical thickness, vertex-wise cortical surface area, cortical thickness, and cortical surface area)^40^. Rs11681562 is an eQTL for multiple genes located in a chromosomal region with high gene density. In this region, our transcriptome-wide and proteome-wide analyses converge on *KHK* as potentially linked to anxiety pathogenesis. This gene is responsible for producing an enzyme involved in fructose metabolism^41^. Animal studies highlighted the potential role of early-life high fructose exposure in long-term depression- and anxiety-like behaviors^42^. *KHK* has also been identified as a PTSD risk locus in transcriptomic and proteomic analyses in the brain and blood^43^. However, we cannot exclude that other genes in this region may contribute to the association observed.

The transcriptome- and proteome-wide analyses converged on five additional loci: *CTNND1*, *CNNM2*, *RAB27B*, *BTN2A1*, and *NEK4*. In both approaches, the strongest statistical evidence was observed for the *CTNND1* gene. This encodes a protein regulating the dendritic spine and synapse development through rho-family GTPases and cadherins^44^. The overexpression of *CTNND1* has been linked to improved memory and reduced anxiety in mice^45^. The *CNNM2* gene, encoding a cyclin protein involved in Mg^2+^ transport, has been associated with impaired neuronal development and epilepsy^46^. *RAB27B* has been identified as a PTSD and depression risk locus in transcriptomic and proteomic analyses in the brain and blood^47^. Reduced *BTN2A1* expression in the placenta was linked to immune system processes in the context of maternal anxiety and depression^48^. *NEK4* has been previously associated with ADHD, BIP, migraines, and cross-disorder psychopathology in genetic and multi-omics analyses^49–52^.

In line with the cross-disorder overlap observed in the single-locus discoveries described above, our genome-wide analyses indicated pleiotropic mechanisms between ANX and other psychiatric disorders. We observed a similar degree of polygenicity and a high genetic correlation between ANX and MDD (rg=0.9), consistent with previous studies^53,54^ and their shared symptomatology^55,56^. However, while 99% of MDD influential variants appear to be shared with ANX, our MixeR analysis suggests that about 10% of the ANX influential variants do not overlap with those of MDD. This may suggest, from a genetic perspective, that MDD is considered an ANX subtype.

LAVA identified several genomic regions with statistical evidence for the local genetic correlation of ANX with multiple psychiatric disorders. In line with the global pleiotropy analysis, ANX shared the most local genetic correlations with MDD. We also observed multiple regions where ANX showed local genetic correlation with two or more mental illnesses. Among them, locus 1719 (chr11:112,755,447-113,889,019) linked ANX to MDD, SCZ, and BIP. Based on data available from the GWAS catalog^57^, over 400 GWS associations involving more than 100 phenotypes have been previously reported in this genomic region (Supplemental Table 16), with the majority related to mental health outcomes, such as anxiety, SCZ, BIP, MDD, PTSD, substance use, and educational attainment. Within this region, we observed the strongest brain-specific TWAS finding, involving the association of ANX with the genetically regulated transcriptomic variation of *DRD2* in the cerebellar hemispheres but not in other brain tissues. This locus is well-known in psychiatric genetics, and considerable research has characterized the function of the encoded dopamine D2 receptor (DR2) protein^58^. In mice, changes in cerebellar D2R levels in Purkinje cells alter sociability and preference for social novelty^59^. The present findings suggest a role for cerebellar D2R in the pathophysiology of human anxiety disorders.

Additionally, we observed a strong genetic overlap between ANX and health-related traits beyond psychiatric disorders. Among the non-psychiatric phenotypes in the 100 highest genetic correlations, we observed several gastrointestinal (43%) and pain-related (17%) phenotypes involving different body areas (e.g., muscle, temporomandibular, and thoracic). Anxiety is highly comorbid with gastrointestinal (GI) disorders. Irritable bowel syndrome has a common genetic basis with anxiety and mood disorders^60,61^. However, the genomic links between anxiety and the broader spectrum of GI disorders are still unclear. A growing body of literature supports the complex interplay between pain and anxiety disorders^62,63^. A recent brain-wide analysis integrating imaging and genetic data highlighted the potential role of pain sensitivity in the pathogenesis of anxiety disorders^64^. The results of our LCV analyses also identified genetic evidence of a causal impact of physical health phenotypes on anxiety disorders, particularly for multiple cardiovascular conditions. This is in line with the known comorbidity between these disorders^65^ and the high anxiety prevalence after cardiac events^66^.

Our common ANX factor demonstrated high loadings to all input datasets (standardized loadings≥0.8), suggesting the generalizability of the results across different cohorts. However, FinnGen and iPSYCH showed more consistent genetic effects than the other samples investigated, with MVP showing the highest degree of phenotypic dilution. One potential explanation is that MVP was the only cohort where anxiety was assessed quantitatively. Those results highlight that more homogeneous and detailed assessments may improve the statistical power and interpretability of genetic studies. Unfortunately, the limited sample size and statistical power in non-EUR populations precluded identifying novel loci in these ancestries. However, we demonstrated evidence that the polygenic risk for ANX detected in EUR is translatable to other population groups. Similar to prior observations^67^, the strength of the cross-ancestry PRS association was proportional to the genetic distance between the population group of the training and target samples. To improve inclusion and equity in anxiety genetics, future studies should prioritize recruiting and assessing diverse cohorts.

In conclusion, this large-scale genome-wide and multi-omics investigation yielded novel insights into the biology of anxiety disorders. We significantly expanded the number of known anxiety risk loci, highlighting the importance of analyzing data from ancestrally diverse participants. Our multi-omics analyses identified novel pathways putatively involved in the pathogenesis of anxiety and refined plausible mechanisms related to previously reported loci. We also disentangled the pleiotropic mechanisms linking ANX to MDD and other psychiatric disorders, reinforcing the importance of studying anxiety disorders in the context of physical health.

## METHODS

### Study Populations

The present study leveraged genome-wide data from six cohorts including AoU^68^, FinnGen^69^, iPSYCH^70^, MVP^10^, PGC^71^, and UKB^28^ (Table 1). This permitted us to investigate a total of 1,266,780 participants with diverse ancestral backgrounds. AoU is a research program aiming to create a representative cohort of the US population to accelerate biomedical research and improve healthcare through precision medicine^68^. Among the samples investigated in the present study, the AoU cohort was the most ancestrally diverse, with information on five genetically inferred population groups (i.e., AFR, AMR, EAS, EUR, and SAS). Details regarding AoU recruitment, assessment, and whole-genome sequencing have been previously described^68^, and the genomic quality control is described at https://support.researchallofus.org/hc/en-us/articles/4617899955092-All-of-Us-Genomic-Quality-Report. In the present study, we analyzed genetic and phenotypic information for 201,334 individuals. The definition of anxiety disorder cases and controls was derived from AoU electronic health records (EHR) using the SNOMED (Systematized Nomenclature of Medicine) classification system^72^. Cases were defined as AoU participants with a lifetime diagnosis of generalized anxiety disorder (SNOMED ID: 434613), panic disorder (SNOMED ID: 4138454), phobic disorder (SNOMED ID: 4304010), chronic stress disorder (SNOMED ID: 436074), acute stress disorder (SNOMED ID: 440083), posttraumatic stress disorder (SNOMED ID: 436676), obsessive-compulsive disorder (SNOMED ID: 440374), or mixed anxiety and depressive disorder (SNOMED ID: 4338031). Any participant without a documented EHR diagnosis falling into the above categories until the maximum follow-up age was considered a control. Ancestry-stratified GWAS was performed using the following quality control criteria: biallelic variants with minor allele frequency >1%, Hardy-Weinberg equilibrium p<10^-6^, call rate >95%, and per-individual genotyping rate >95%. A PLINK 2^73^ logistic regression model estimated genetic associations, including covariates for sex, age, and the first ten within-ancestry principal components.

FinnGen is a project focused on developing a large Finnish cohort combining genotype data from biobanks and digital health record data from health registries^27^. In the present study, we used genome-wide association statistics generated from Release 9 (May 11, 2023). A detailed description of FinnGen data is available at https://finngen.gitbook.io/documentation/v/r9/. The FinnGen GWAS was performed using REGENIE^74^ including covariates for age, sex, the top-ten within-ancestry PCs, and genotyping batch. In our analysis, we used GWAS data generated from 40,191 cases and 277,526 controls of EUR descent for the phenotype “anxiety disorders” (KRA_PSY_ANXIETY), defined based on International Classification of Diseases (ICD-10 F40-F48; ICD-9 300.0-300.3, 300.6-300.9,3078A, 309; ICD-8 300.0-300.2, 30030, 300.5-300.9, 305, 30680, 30799). Details regarding the KRA_PSY_ANXIETY definition are available at https://risteys.finregistry.fi/endpoints/KRA_PSY_ANXIETY.

iPSYCH is a large Danish population-based cohort aimed at unraveling the genetic and environmental architecture of severe mental disorders^75^. In the present study, we used genome-wide association statistics generated from the analysis of 12,655 cases and 19,225 controls of EUR descent^70^. Cases were defined as individuals with an anxiety and stress-related diagnosis according to the ICD-10 F40.0-F41.9 and F43.0-F43.9. codes. Controls included individuals without any ICD-10 diagnoses of anxiety, stress-related disorders, or mood disorders. Genetic associations were estimated using logistic regression models with the imputed marker dosages using the first four principal components as ancestry covariates. The analysis was stratified by genotyping batch, and the results were meta-analyzed using an inverse variance–weighted fixed-effect models.

MVP is a biobank funded by the US Department of Veterans Affairs to understand how genes, lifestyle, military experiences, and exposures affect health and wellness^76^. The GWA statistics used in the present study were generated from an analysis of 61,796 and 241,541 participants of AFR and EUR descent, respectively^10^. The anxiety phenotype was the total score of the generalized anxiety disorder 2-item scale, which ranged from 0 to 6. The genetic association analysis was performed using a PLINK 2^73^ logistic regression model with covariates for age, sex, and the first ten within-ancestry principal components.

PGC genome-wide association statistics were derived from a meta-analysis conducted by the Anxiety NeuroGenetics STudy (ANGST)^71^. Cases were defined as individuals with diagnoses of generalized anxiety disorder, panic disorder, social phobia, agoraphobia, or specific phobias^71^. A total of 5,761 cases and 11,765 controls of EUR descent were included. The GWAS was performed using a logistic regression model with covariates for sex, age, and the first ten within-ancestry principal components.

UKB is a large population-based study that collected information regarding over 500,000 participants^28^. GWA statistics used in the present study were obtained from the Pan-UKB analysis. Details regarding the quality control, ancestry assignment, and statistical analyses are available at https://pan.ukbb.broadinstitute.org/. Briefly, ancestry-stratified GWASs were performed using the Scalable and Accurate Implementation of Generalized (SAIGE)^77^ mixed models, including a kinship matrix as a random effect and the remaining covariates (i.e., top-10 within-ancestry PC, sex, age, age^2^, sex×age, and sex×age^2^) as fixed effects. We analyzed UKB a total of 10,751 cases and 383,235 controls of AFR, EUR, and SAS descents for the phecode 300 “Anxiety, dissociative and somatoform disorders”.

### Phenotype Definition Comparisons

Because of the different anxiety definitions, we compared their genetic architecture using two methods. LD Score regression^78^ was used to estimate their SNP-based heritability and their pairwise genetic correlations. LD scores were calculated using HapMap 3 variants and the 1000 Genomes Project reference populations corresponding to each ancestry group^79^. Phenotype dilution among anxiety definitions was quantified using PheMED^14^, considering the FinnGen cohort as the reference sample because it showed the highest SNP-based heritability z-score. Because of the lack of significant SNP-based heritability in non-EUR samples, genetic correlation, and phenotype dilution analyses were performed only in EUR datasets.

### Ancestry-Specific and Cross-Ancestry Meta-Analyses and Conditional Analysis

Due to moderate genetic correlation (median rg=0.72) and statistically significant phenotype dilution among anxiety phenotypes, we combined the EUR GWASs using the gSEM approach^15^. As recommended^80^, we used the effective sample size for each EUR anxiety GWAS. For binary definitions, this was calculated as 4/(1/*N_cases_*+1/*N_controls_*). For quantitative phenotypes, we used the total sample size. The effective sample size of the common ANX factor obtained from the gSEM analysis was estimated as *mean*(1/(2×*MAF*×(1–*MAF*)×*SE*^2^), where MAF is the minor allele frequency and SE is the standard error. Due to the limited sample sizes, we could not use gSEM to combine genome-wide information available for AFR (AoU, UKB, and MVP) and SAS (AoU and UKB). Thus, we performed ancestry-specific meta-analyses using the sample-size weighted approach available in METAL^18^. To present ancestry-specific effects with the same statistics, AFR and SAS z-scores obtained from the sample-size weighted approach were converted to betas using the following formula: beta = *z*⁄*sqr*t(2 ×*p* × (1 – *p*) × (*n*+ *z*^2^), where p is the effect allele frequency and n is the sample size. The sample-size weighted approach, using effective sample sizes, was also used to perform the cross-ancestry meta-analysis. Independent associations were identified by clumping GWS associations considering LD r^2^<0.1. To identify secondary associations accounting for the primary GWS signals, we applied the stepwise model selection procedure available in the COJO approach implemented in the GCTA package^17,81^. Among the GWS associations identified by the conditional analysis, we considered only those with LD r^2^<0.8 with respect to the corresponding index GWS variant.

### Cross-Ancestry Polygenic Risk Scoring

To investigate how anxiety polygenic risk translates across ancestry groups, we derived PRS from the EUR ANX factor and tested them in other populations available from the AoU cohort (i.e., AFR, AMR, EAS, and SAS). To compare within-ancestry to cross-ancestry PRS associations, we derived a PRS from the EUR ANX factor excluding AoU-EUR and used the latter as the EUR target sample. Posterior variant-level effect sizes of the EUR ANX factor were computed using PRS-CS^82^ and the 1000 Genomes European population as LD reference. ANX PRS were computed using PLINK^73^. We tested the quintiles of PRS distribution using the first quintile as reference with logistic regression models available in the *stats* R package in AoU population groups.

### Heritability Analyses

LDSC^78^ was used to estimate the SNP-based heritability of anxiety leveraging ancestry-specific genome-wide association statistics using the methods described above. For the EUR ANX factor, we also performed a partitioned heritability analysis^83^, testing the enrichment of 205 tissue- and cell-type specific transcriptomic profiles. These annotations were derived from GTEx V8^22^ (53 tissues) and the Franke lab^84,85^ (152 tissues/cell types).

### Transcriptome- and Proteome-Wide Association Studies

To understand the role of genetically regulated transcriptomic and proteomic variation in anxiety pathogenesis, we performed transcriptome-wide and proteome-wide association analyses. A brain-specific transcriptome-wide investigation was conducted using S-PrediXcan^21^ to integrate EUR-ANX genome-wide association statistics with GTEx eQTL data available for 13 brain tissues. A Bonferroni correction accounting for the number of tests performed (N=165,710, p<3×10^-7^) was applied to define the statistically significant associations. To explore cross-tissue transcriptomic regulation, we performed a multi-tissue transcriptome-wide association analysis using S-MultiXcan^23^. This permitted us to boost the statistical power of the association analysis via a joint multi-tissue analysis accounting for transcriptomic correlation across the 49 GTEx tissues tested. Multi-tissue genetically regulated transcriptomic associations were defined after Bonferroni correction accounting for the number of the genes tested (N=22,045; p<2.27×10^-6^).

Proteome-wide associations were estimated using the FUSION approach^24^ to integrate EUR-ANX GWAS with dlPFC pQTLs available from Religious Orders Study and Memory and Aging Project (ROSMAP)^86^ and the Banner Sun Health Research Institute (BSHRI)^87^. Analyses were performed separately in two pQTL datasets and the results were meta-analyzed using an inverse-variance weighted approach available in METAL^18^. Bonferroni correction accounting for the number of genes tested (N=1,629; p<3.07×10^-5^) was applied to define proteome-wide significant genes. To understand their implications on synaptic localization and function, these loci were analyzed using SynGO^88^, an online knowledge base that organizes research on synaptic proteins using GO annotations. SynGO enrichments were estimated for each subset of cellular components and biological processes by a one-sided Fisher exact test.

### Pleiotropy Analyses

We used multiple methods and data sources to estimate the pleiotropy between EUR ANX and human traits and diseases. Initially, we focused on understanding the shared genetic mechanisms of ANX with other psychiatric disorders previously investigated using GWAS, including ADHD^89^, AN^90^, ASD^91^, BIP^92^, MDD^93^, OCD^94^, PTSD^95^, SCZ, and TS^96^. After excluding the MHC region (chr6:26,000,000-34,000,000), we applied mixture models available from MixeR^25^ to estimate the number of influential variants ANX shares with other psychiatric disorders. We then applied LAVA^26^ to estimate local genetic correlations of ANX with mental illnesses. Because we observed local genetic correlation with multiple psychiatric disorders in the same genomic regions, we used LAVA multivariate models^26^ to dissect whether the observed local genetic correlations were common or distinct among the implicated disorders. For MixeR and LAVA analyses, European populations available from 1000 Genomes Project Phase 3 were used as LD reference.

To assess ANX pleiotropic mechanisms across the human phenotypic spectrum, we performed an LDSC genetic correlation analysis^78^ for 11,175 phenotypes available from UKB (details available at https://pan.ukbb.broadinstitute.org/), FinnGen (Release 9, details available at https://finngen.gitbook.io/documentation/v/r9/), and MVP^29^. Bonferroni correction accounting for the number of tests performed was applied to define statistically significant genetic correlations (p<4.47×10^-6^). To investigate the potential causality underlying the observed genetic correlations, we conducted a phenome-wide LCV^30^ analysis using FinnGen, UKB, and MVP data. This was performed using LD scores calculated from European populations available from the 1000 Genomes Project Phase 3. LCV permitted us to calculate gcp estimates. Positive and negative gcp values reflect the direction of the putative causal effect (i.e., phenotype #1 → phenotype #2 and phenotype #2 → phenotype #1, respectively). In the present study, positive gcp values correspond to the effect of a phenotype on ANX. Information regarding the sign of the LCV effect is determined by the LCV rho statistics: rho>0 corresponds to a positive effect, while rho<0 corresponds to a negative effect.

## Data Availability

All data produced in the present study are available upon reasonable request to the authors

## ACKNOWLEDGMENTS

This study was supported by grants from the National Institutes of Health (RF1 MH132337, R33 DA047527, and K99 AG078503), One Mind, the Alzheimer’s Association (Research Fellowship AARF-22-967171), the American Foundation for Suicide Prevention (PDF-1-022-21), Horizon 2020 (Marie Sklodowska-Curie Individual Fellowship 101028810), University of Bergen (International Training Grant), and the Yale Franke Program in Science and Humanities. We also acknowledge the contribution of the participants and the investigators involved in the UK Biobank, the FinnGen Project, the Million Veteran Program, the All of Us Research Program, the iPSYCH study, and the Psychiatric Genomics Consortium. The All of Us Research Program is supported by the National Institutes of Health, Office of the Director: Regional Medical Centers: 1 OT2 OD026549; 1 OT2 OD026554; 1 OT2 OD026557; 1 OT2 OD026556; 1 OT2 OD026550; 1 OT2 OD 026552; 1 OT2 OD026553; 1 OT2 OD026548; 1 OT2 OD026551; 1 OT2 OD026555; IAA #: AOD 16037; Federally Qualified Health Centers: HHSN 263201600085U; Data and Research Center: 5 U2C OD023196; Biobank: 1 U24 OD023121; The Participant Center: U24 OD023176; Participant Technology Systems Center: 1 U24 OD023163; Communications and Engagement: 3 OT2 OD023205; 3 OT2 OD023206; and Community Partners: 1 OT2 OD025277; 3 OT2 OD025315; 1 OT2 OD025337; 1 OT2 OD025276.

