## Supplementary Figures for "Gene Discovery and Biological Insights into Anxiety Disorders from a Multi-Ancestry Genome-wide Association Study of >1.2 Million Participants"

**Supplementary Figure 1**. Phenotypic dilution among anxiety phenotypes assessed in EUR participants. The asterisk denotes cohorts with statistically significant dilution statistics when compared to the FinnGen sample. AoU: All of Us Research Program; MVP: Million Veterans Program; PGC: Psychiatric Genomics Consortium; UKB: UK Biobank.

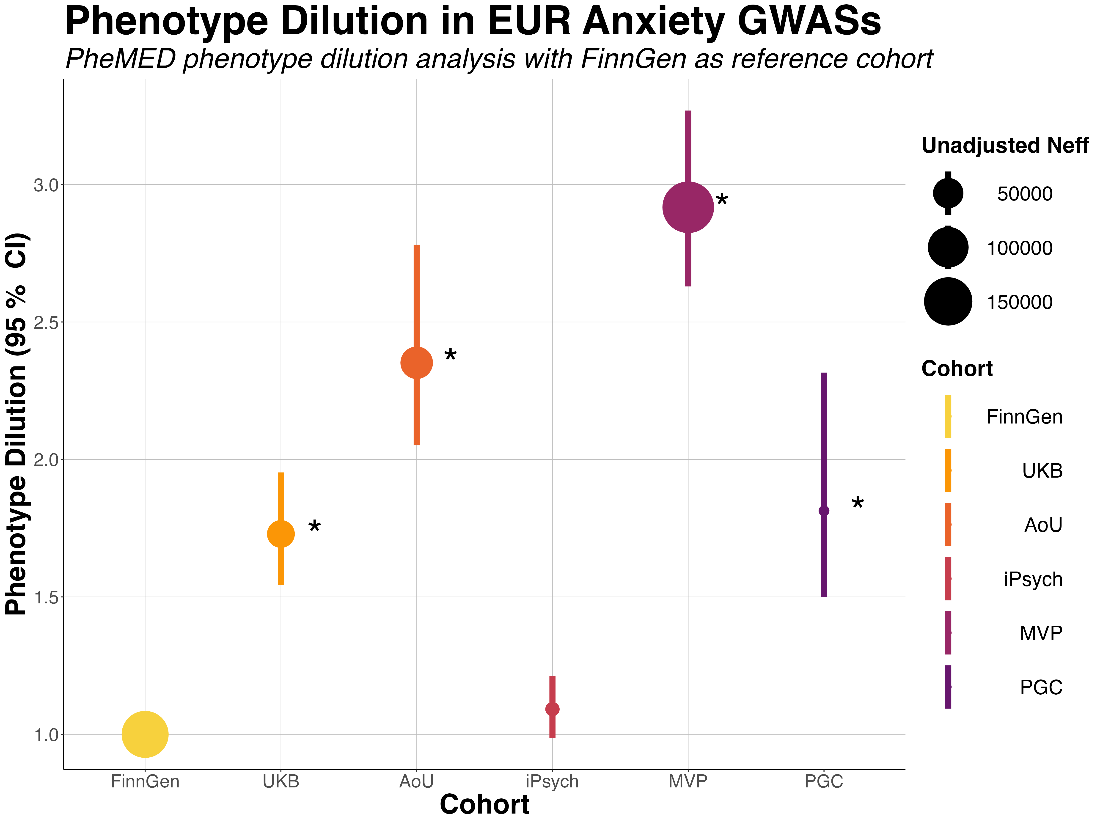

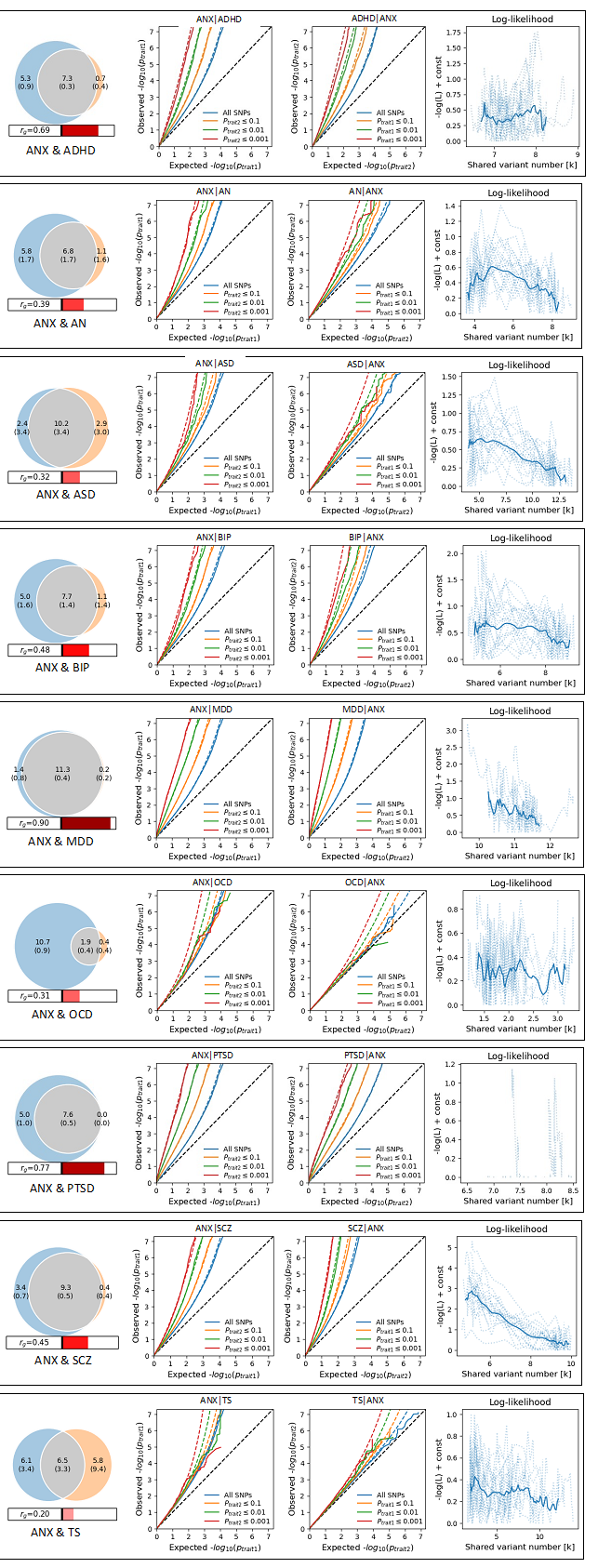

**Supplementary Figure 2:** Venn Diagrams of the number of shared and distinct causal SNPs of ANX with nine psychiatric disorders given by bivariate mixture models.

**Supplementary Figure 3:** Local genetic correlations in locus 1719

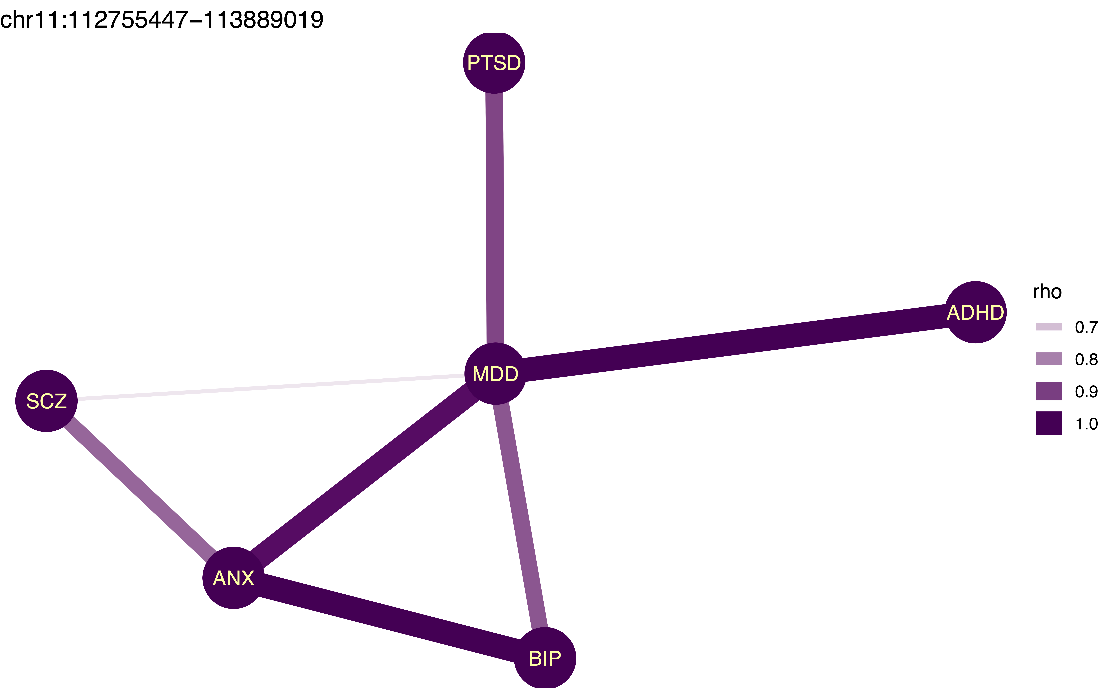
